# Chinese herbal medicine for varicocele in subfertile men A protocol for systematic review and meta-analysis

**DOI:** 10.1101/2023.01.09.23284339

**Authors:** Wenting Tong, Zhimin Zhao, Xuchong Tu, Shan He, Yongxing Wang, Ming Chen, Hui Zhang

## Abstract

**Background:** The purpose of this protocol is to provide a updated systematic review and meta-analysis to prove the effectiveness and safety of Chinese herbal medicine in the treatment for the patients with varicocele.

**Method:** This protocol conforms to the Preferred Reporting Items for Systematic Review and Meta-Analysis Protocols (PRISMA-P) and the recommendations of the Cochrane Collaboration Handbook. We selected qualified studies published as of May 1, 2022, and systematically searched 6 English database (Embase, Pubmed, Scopus, Web of Science, Cochrane Central of Controlled Trials (CENTRAL), and Clinicaltrials.gov) and 5 Chinese database (China National Knowledge Infrastructure (CNKI), VIP Database for Chinese Technical Periodicals, Wanfandata, SinoMed, and Chinese Clinical Trial Registry). At the same time, relevant reviews and a list of references included in the study were retrieved, and Epistemonikos.org, ISI Web of Science and OpenGrey were manually searched to screen any other studies not included in the previous search. There will be no language restrictions. The inclusion criteria were clinical randomized controlled trial (RCT) involving the use of traditional Chinese medicine in the treatment of varicocele. The main results were fertility rate, adverse events, semen quality and scrotal pain score after 3 months, 6 months and 1 year follow-up. Bias analysis and evaluation will be performed based on risk of bias (ROB) assessment tool provided by the Cochrane Handbook. And use GRADEpro GDT to grade, evaluate and score the quality of the evidence. Heterogeneity will be judged by I^2^ value. At the same time, report bias assessment, subgroup analysis and sensitivity analysis were carried out. According to the Cochrane Manual of Systematic Evaluation of Interventions (Higgins 2011), if the data showed sufficiently high quality and some degree of similarity, we included the data for the meta-analysis. For dichotomy data, we selected an effect scale relative risk (RR) represented by a 95% confidence interval (CI). The continuous data is expressed as mean difference (MD) or standardized mean difference (SMD).

**Result:** This study will provide high-quality evidence for the efficacy and safety of Chinese herbal medicine in the treatment of varicocele in subfertile men.

**Conclusion:** This study will provide an effective and safe choice for Chinese herbal medicine to improve the fertility of patients with varicocele.

**Ethics:** The data of this study are based on published studies and do not require additional ethical approval. We will publish our findings through peer-reviewed journals.

PROSPERO database registration number: CRD42022331218

## 1 Introduction

Varicocele is closely related to male infertility [1, 2] and is considered to be the most common correctable male infertility disease [3-5], about 15% of the normal population and 35% of infertile patients suffer from VC [5]. The pathogenesis of VC is not clear. At present, the mainstream view is the increase of local temperature in the scrotum [6], oxidative stress [7], and the reflux of metabolites in kidney and adrenal gland [8] lead to decreased spermatogenic function and poor sperm quality. Although some patients with varicocele can give birth normally without intervention, most patients still need active intervention to achieve better results [9]. At present, surgical treatment is widely used in patients with abnormal semen parameters [10]. However, surgical repair is not always accompanied by an improvement in sperm quality [11] and an increase in pregnancy rate [12].

Therefore, some patients and doctors try to seek adjuvant treatment of Chinese herbal medicine to improve the curative effect. Although some basic studies have confirmed that the use of traditional Chinese medicine can improve sperm quality through antioxidant and anti-inflammatory effects [13-16], the lack of clinical evidence still makes the application of Chinese herbal medicine in the shadow. Before this study, some researchers tried to make a meta-analysis of the effects of Chinese herbal medicine, but did not get a credible result [17]. Therefore, on the basis of previous studies, this study will further include new studies, optimize the retrieval strategy, and systematically evaluate the improvement effect and safety of Traditional Chinese medicine on the fertility of patients with varicocele, so as to provide evidence-based medical basis for clinical medication.

## 2 Methods and analysis

The study was conducted following the guidelines of the Preferred Reporting Items for Systematic Review and Meta-analysis Protocol (PRISMA-P) This study protocols have been funded through a protocol registry. This protocol of the systematic review has been registered in the PROSPERO database (Registration number: CRD42022331218).

### 2.1 Inclusion criteria

#### 2.1.1 Types of participants

This study includes participants who having had normal sex (or married) for more than 2 years but did not make the spouse pregnant. Participants who were infertile due to immunization problems, female fertility problems, or other causes were excluded.

#### 2.1.2 Types of interventions

Various forms of formula, including decoction, pill, electuary, with or without other therapeutic interventions, can be included and meets the following criteria.

1. Traditional Chinese Herb versus no treatment/any treatment.
2. Traditional Chinese Herb combined with other therapies versus the same other therapies.

For groups that use the same prescription but have multiple doses, their results will be combined. If there are multiple dose groups, first determine whether the same prescription has a clear target dose (standard dose). If there is a definite target dose, we only need to extract the data of the target dose group and combine the target dose group with the control group for meta-analysis. If there is no clear target dose, it is necessary to compare whether the dose groups included in the literature are consistent. If there is consistency, the data of each dose group are extracted independently and meta-analysis. If there is no consistency, the data of multiple different dose groups need to be merged, and the mean and SD of multiple dose combinations can be calculated according to the following formula. Suppose there are two different dose groups, the sample size of dose group A is N_1_, the mean is M_1_, and the standard deviation of SD_1_; dose group B is N_2_, the mean is M_2_, and the standard deviation is SD_2_. The combined sample size is N= N_1_+ N_2_, the mean is M = (N_1_ M_1_+ N_2_ M_2_) / (N_1_+ N_2_), and the SD is:

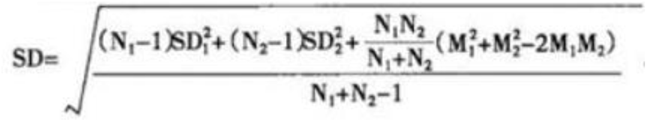

Assuming that there are several different dose groups, according to the above formula, the data of two dose groups are merged first, then the data obtained are combined with the third dose group, and so on.

### 2.1.3 Types of studies

This study is intended to include a randomized controlled clinical trial (RCT) involving the use of Chinese herbal medicine in the treatment of varicocele. Animal experiments, case reports and non-randomized controlled trials were excluded. And there will be no additional restrictions on the language.

#### 2.1.4 Types of outcomes

According to the recommendations of the guidelines and previous studies[18-20]. The following main outcome indicators were set.

##### 2.1.4.1 Primary outcomes

a. Fertility rate at 3 months, 6 months, and 1 year after follow-up
b. Adverse events

##### 2.1.4.2 Secondary outcomes

a. Semen quality
b. Scrotal pain score

### 2.2 Data sources and search methods

#### 2.2.1 Electronic searches

We systematically searched 6 English database (Embase, Pubmed, Scopus, Web of Science, Cochrane Central of Controlled Trials (CENTRAL), and Clinicaltrials.gov) and 5 Chinese database (China National Knowledge Infrastructure (CNKI), VIP Database for Chinese Technical Periodicals, Wanfandata, SinoMed, and Chinese Clinical Trial Registry). In addition, we will retrieve relevant reviews and a list of references included in the study, and manually search Epistemonikos.org, ISI Web of Science and OpenGrey to find any other studies not included in the previous search.

We selected qualified studies published as of May 1, 2022. The combination of subject terms and free terms is used as the retrieval strategy. The retrieval strategy is decided by all the participants. The main search terms are varicocele, varicocelectomy and Chinese herbal medicine. Take Embase as an example, the retrieval strategy is shown in Table 1. And adjust the specific database accordingly.

### 2.3 Data collection and export

All the literatures were imported into Endnote X9 software (Thomson Research Soft, Stanford, Connecticut). First of all, duplicate documents are eliminated through the duplicate checking function of endnote, and then two independent reviewer (Wenting Tong and Zhimin Zhao) read titles and abstracts to eliminate unqualified documents, and then conduct full-text screening of the remaining documents to determine whether they meet the inclusion requirements, and seek the help of language experts for studies that need translation. Finally, the two reviewer compare the screening process for consistency according to the PRISMA flowchart. Any differences during the screening process will be agreed upon through discussions between the reviewers or by seeking the advice of a third reviewer (Hui Zhang). The selection process is recorded in the PRISMA flow chart [21].

### 2.4 Data extraction and analysis

Develop an excel data extraction table and extract data independently from the included qualified studies by two independent reviewer. Any differences will be settled by consensus. Any differences in the extraction process will be agreed upon through discussions between the two parties or seeking the advice of a third party (Hui Zhang). The main data extracted are as follows:

Title, author, year, source of funding, sample size, age, duration of disease, specific interventions, Fertility rate (if applicable), how to confirm pregnancy, semen quality (if applicable), scrotal pain score (if applicable), adverse events. And the randomization method of the study, the specific blind method, the nature of the study, the follow-up time, and record the basic situation of the woman, such as the age of the partner and whether to exclude the infertility caused by the female factor (if applicable). If the data are missing, try to contact the lead author of the study.

### 2.5 Assessment of risk of bias in the included studies

The two researchers will conduct bias analysis and assessment based on risk of bias (ROB) assessment tool provided by the Cochrane Handbook [22]: selection bias (random sequence generation and allocation concealment); performance bias (blinding of participants and personnel); detection bias (blinding of outcome assessors); attrition bias (incomplete outcome data); reporting bias (selective reporting),and other bias. Evaluate and fully describe the basis of judgment through the three grades of “low bias”, “high bias” and “ambiguity bias”. Any differences in the screening process shall be discussed by both parties or seek third parties (Hui Zhang).

### 2.6 Assessment of heterogeneity

The heterogeneity of this study will be judged by the I^2^ value. If I^2^ < 50%, P > 0.1, we believe that there is no statistical heterogeneity among the included studies, and the fixed effect model (FEM) will be used for meta analysis. If there is obvious heterogeneity (I^2^ < 50%, P > 0.1), we use the random effect model (REM) for meta analysis, or consider a descriptive analysis without merging the results.

### 2.7 Assessment of reporting biases

For publication bias, funnel chart and Egger linear regression analysis to test whether there is publication bias were used if the number of studies included was more than 10. The report bias will be done through R software.

### 2.8 Data synthesis

According to the Cochrane Manual of Systematic Evaluation of Interventions (Higgins 2011), if the data showed sufficiently high quality and some degree of similarity, we included the data in the meta-analysis. For dichotomy data, we selected an effect scale relative risk (RR) represented by a 95% confidence interval (CI). The continuous data is expressed as mean difference (MD) or standardized mean difference (SMD). We plan to compare the baseline to the changes after the intervention rather than individual final measurements. According to the heterogeneity test results, we choose fixed effect model or random effect model. According to the specific situation, subgroup analysis and meta regression were carried out. If it is not suitable for merger analysis, descriptive analysis is used. If there is a lack of key data in the study, we will try to contact the main authors of the study to obtain the missing data. If the above methods can’t help us to obtain data, we will take the Markov Chain Monte Carlo (MCMC) stochastic simulation method as the guidance, based on the multiple filling method to deal with the missing values, in order to fit the missing data to the maximum extent.

### 2.9 Subgroup analysis

If non-statistical heterogeneity is detected, the factors that may cause heterogeneity are grouped to explore possible explanations, such as the course of disease, the grading of varicocele, the dosage of Chinese herbal medicine, the mode of combined intervention and so on. If necessary, the study of subclinical varicocele is distinguished.

### 2.10 Sensitivity analysis

We conduct sensitivity analysis to determine the stability and reliability of the results of this study, and sensitivity analysis will be carried out through R software.

### 2.11 Grading the quality of evidence

We use Grading of Recommendations, Assessment, Development and Evaluation (GRADE) tool to evaluate the quality of the evidence. The comparison will be evaluated by two reviewers based on recommended ratings, and any inconsistencies in the assessment will be decided by a third reviewer.

## 3. Discussion

Varicocele surgery is often used in patients who expect to improve sperm parameters, but successful surgery does not means acceptable results. Apart from the improvement in the reproductive capacity of some patients [23], many patients did not achieved significant benefits [1]. As a supplementary treatment of surgery, Chinese herbal medicine is widely used in the treatment of varicocele. However, because the shortage of existing research, the role of Chinese herbal medicine in the treatment is still confusing, and it is worth more discussion on related issues. In addition, the ingredients of Chinese herbal medicine decoction are complex and may contain ingredients that cause side effects. Therefore, the side effects in the use of Chinese herbal medicine are also the issues we need to concern.

We tried to evaluate specific prescriptions in this study, however, because the regulation of drugs in traditional Chinese medicine is reflected in the dose of individual drugs in the prescription, and that each individual has a unique physique, based on the different understanding of the theory of traditional Chinese medicine, it is difficult to use the same prescription for comprehensive analysis. Therefore, we regard traditional Chinese medicine as a whole intervention and look for evidence-based medicine that Chinese herbal medicine prescription plays a role in the adjuvant treatment of varicocele, just like the research related to acupuncture [24, 25].

## Data Availability

Data sharing not applicable to this article as no datasets were generated or analyzed during
the current study.

## Declarations

### Ethics approval and consent to participate

Not applicable.

### Consent for publication

Not applicable.

### Availability of data and material

Data sharing not applicable to this article as no datasets were generated or analyzed during the current study.

### Authors’ contributions

Conceptualization: Wenting Tong and Shan He; Data curation: Wenting Tong and Zhimin Zhao; Funding acquisition: Hui Zhang; Methodology: Wenting Tong and Yongxing Wang; Project administration: Hui Zhang and Ming Chen; Supervision: Xuchong Tu; Validation: Zhimin Zhao and Yongxing Wang; Visualization: Shan He and Ming Chen; Writing – original draft: Wenting Tong and Xuchong Tu; Writing – review & editing: Wenting Tong, Zhimin Zhao and Hui Zhang.

## Acknowledgements

Not applicable.

---

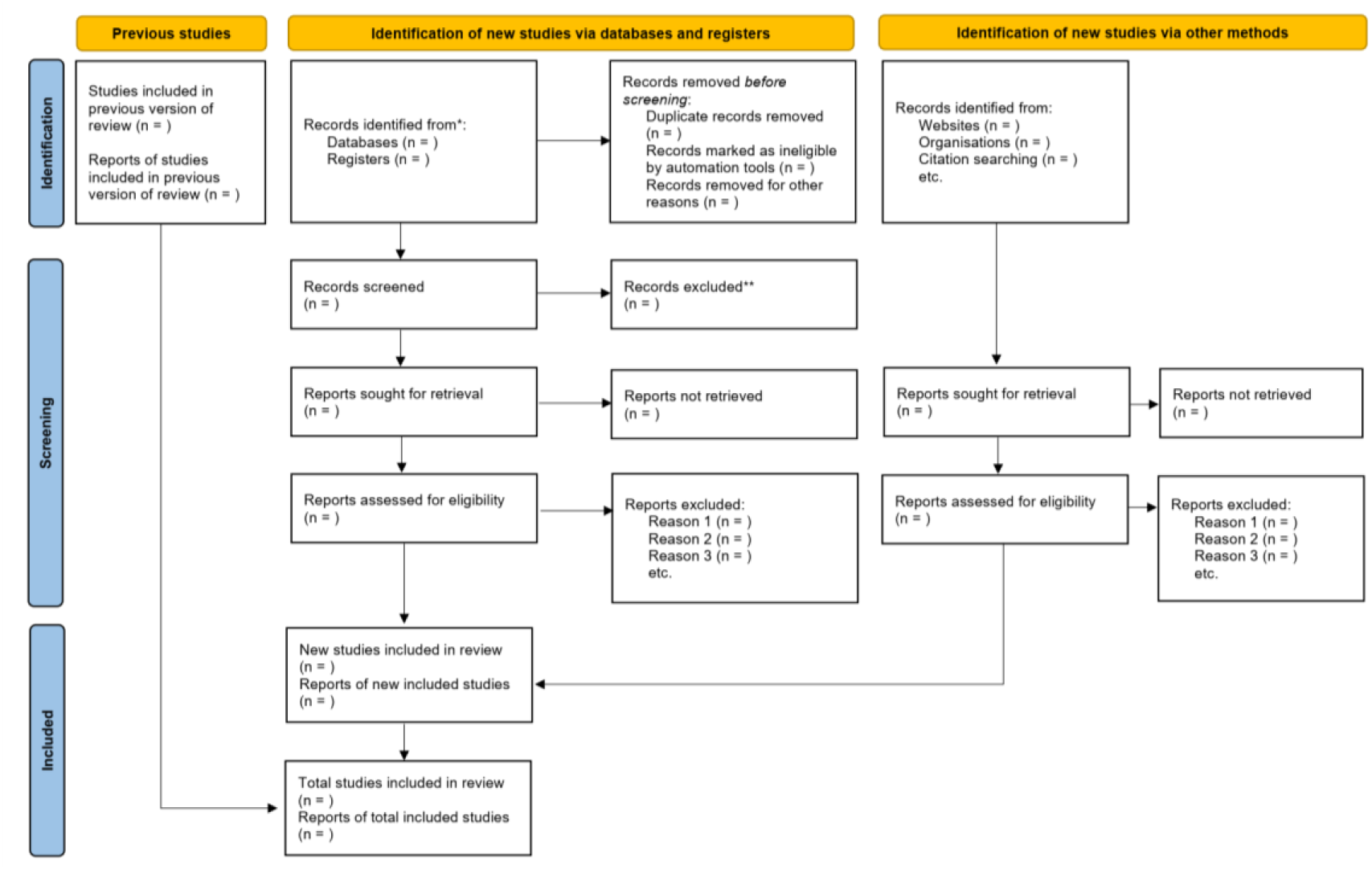

